# Risk of COVID-19 infection and work place exposure of front-line mass media professionals

**DOI:** 10.1101/2021.05.06.21256773

**Authors:** Sarabon Tahura, Bilkis Banu, Nasrin Akter, Sarder Mahmud Hossain, Rashidul Alam Mahumud, Md Rishad Ahmed

## Abstract

**Introduction:** Mass media plays a crucial role in creating awareness and knowledge sharing in this Corona virus disease 2019 (COVID-19) pandemic. However, the risk of exposure and extent of COVID-19 infection among media professional are less elucidated yet. Therefore, this study was intended to investigate the workplace-related risk of COVID-19 exposure and the association between exposure to COVID-19 and participant’s characteristics, including various forms of respiratory protection for mass-media professionals.

**Methods:** This closed web-based cross-sectional survey was conducted among 199 mass-media professionals in Bangladesh by snowball sampling approach. A multivariate logistic regression model was used for the analytical exploration. Adjusted and Unadjusted Odds Ratio (OR) with 95% confidence intervals (95% CI) were calculated for the specified exposures. Chi-square test was used to observe the association. Ethical issues were maintained according to the guidance of the declaration of the Helsinki.

**Results:** Of all, 39.2% of mass-media professionals were tested positive for COVID-19, whereas 6% of symptomatic or suspected participants did not do the test. Mass media professionals who worked in electronic media reported more COVID-19 infection (adjusted odds ratio, AOR= 6.25; 95% Confidence interval: Lower limit 1.43, upper limit 27.43; P =0.02). However, no significant relationship was found between the type of job role and COVID-19 infection. Furthermore, infected colleagues (OR/P=1.92/0.04) were identified as significant contact of acquiring infection. However, the study result showed that reused/new medical mask, homemade/cloth-made mask (vs. use of respirator mask) was not significantly (p=0.82) associated with mass media professional’s infection.

**Conclusions:** Professionals working in electronic media were at higher risk of being infected by COVID-19 and mostly acquired from infected colleagues. Using a respirator mask was not associated with a lower risk of test positive infection in mass media professionals. This study will aid the policy maker and public health authorities during the COVID-19 pandemic to make proper implementation strategies.

## Introduction

In early December 2019, a cluster of pneumonia cases of unknown cause emerged in Wuhan, Hubei province, China [1]. Chinese Center for Disease Control and Prevention (China CDC) confirmed as the cause of this disease is a novel member of enveloped RNA coronavirus [2–4]. On February 11, 2020, World Health Organization (WHO) named the illness associated with 2019-nCoV as the 2019 corona virus disease (COVID-19) [5]. On March 11, 2020, the WHO declared Covid-19 as a pandemic [6].

Transmission of the virus mostly occurred by respiratory droplets produced by sneezing, coughing [7] and by fomites in the immediate environment around the infected person [8]. Therefore, both direct contact with infected people and indirect contact with inanimate surfaces can acts as a source of infection. So, controlling strategy against spread of the virus are thought to be the most effective measure yet. On the other hand, fear and anxiety about the COVID-19 pandemic are causing overwhelming stress for everyone [9]. Conversely, ample misinformation and conspiracy theories about the origin, prevention, diagnosis, and treatment of the disease have been spread through social media and text messaging [10]. COVID-19 epidemic is showing the critical impact and role of information diffusion among the general population. In this crisis, print and electronic broadcast media across the world are working not only to cover the pandemic, providing health updates but also plays an important role to create awareness among the mass population [11].

In Bangladesh, the first three Covid-19 cases were reported on March 8, 2020, by the Institute of Epidemiology, Disease Control and Research (IEDCR) [12]. Like the other countries, Bangladesh’s print and electronic media professionals are also putting in efforts to take up people’s voice with governments regarding the management of COVID-19 outbreak, fill information gaps and counter misinformation, and create awareness of the contagion among people. But, mass media professionals, especially video journalists and news reporters, may not maintain physical distancing when they are on duties, as sometimes they have largely crowded places or throng together while covering events or press briefings. Thus, since the emergence of the COVID-19 pandemic, work settings have become challenging and appear at the front line for mass-media professionals. However, a substantial outbreak could dramatically disrupt their professional duties and become a threat to public health. Balancing the duty to work with the fear and anxiety of being infected and transmitted infection from the workplace is a core issue for front-line employees during a pandemic [13]. And on the other hand, this feeling of insecurity and fear among employees aggravate by insufficient information and suboptimal protective management of the workplace’s response to the crisis [14]. So, it is imperative to determine which work settings and exposures place mass-media professionals at greater risk and what protective measures reduces their risk. There have been no studies investigating their infection status and workplace-related risk to the best of our knowledge.

The aim of this study was to investigate the workplace-related risk of COVID-19 exposure and the association between exposure to COVID-19 and participant’s characteristics, including co-morbid conditions, various forms of respiratory protection for mass-media professionals. The findings of this study would generate baseline information related to risk of mass media professionals, and will aid the policy maker to make proper implementation measures.

## Methods

### Settings and Participants

A cross-sectional study was conducted among mass media professionals in Bangladesh, who were actively working from May 2020 to June 2020 to delineate the disease transmission among Bangladesh’s electronic and print media professionals.

A total of 220 respondents have participated in this survey. However, 21 samples were excluded from the analysis due to insufficient information. Finally, a total of 199 respondents were included in this current analysis, who were working in different print (newspapers), broadcast (Television channels), and online mass media available in Bangladesh. Data were collected using the exponential non-discriminative snowball sampling technique. The first respondent was recruited to the sample group provides multiple referrals. Each new referral was explored until primary data from a sufficient number of samples were collected. Any print or electronic media, who had been diagnosed with RT polymerase chain reaction (PCR) test or who had experienced an illness suspicious for COVID-19 during the pandemic without test confirmation and the media professionals who worked during the pandemic in the same settings were also included in this study.

Initially, we assumed a potential standard sample size 384 by using the formula “n=‘Z^2^pq/d^2^” where Z (standard normal deviate) was considered as 1.96; p (proportion of media professionals) was unknown and was considered as 0.50 and margin of error was considered as 0.05. However, online and snowball sampling strategies having some high non-response rates.

### Procedure

Data were gathered by a self-administered method by using a structured and anonymous online questionnaire. Due to the ongoing lockdown adopted by the country officials in Bangladesh, a physical and paper-based questionnaire was not feasible. Thus, to collect the data email and social media platforms such as WhatsApp and Facebook messenger were used concurrently. The web link of the online survey was ‘http://covid19.dreamhomebd.net/’, which took only 8 to 10 minutes to complete by the respondents.

### Measures

The online questionnaire was developed by using Google forms. Prior to conduct the survey, the questionnaire was pretested among 10 respondents. Experience from the pre-testing were adjusted during finalization of the questionnaire. The questionnaire comprised of several segments: (i) Identification of COVID-19 patients among the respondents (ii) Demography of respondents: age; gender (iii) Clinical and health indicators: physical symptoms; comorbidities/ chronic medical conditions; tobacco smoking status; contact tracing and seeking the maximum medical care support (iv) Media workplace information: media type; job role; types of masks used (v) Exposure status: exposure pattern within the workplace and outside setting; status of exposure to COVID-19 test positive person and a person under investigation of COVID-19.

### Data analysis

Data were entered, checked for quality, and analyzed utilizing the Statistical Package for the Social Sciences (SPSS) software, v.22. To summarize the obtained data, study characteristics were subjected to descriptive statistics (frequency and proportions). A multinomial logistic regression analysis was performed, including pre-specified confounders age, gender, symptoms, co-morbidities, tobacco smoking status, contact tracing, seeking of maximum medical care support, media type; job role; types of masks used. Adjusted and Unadjusted Odds Ratio (OR) with 95% confidence intervals (95% CI) with respect to COVID-19 status, i.e., test positive and suspected test negative and healthy (Dummied the three groups: suspected with test negative, suspected without test and healthy or test not done in one group) were calculated for the specified exposures. A Chi-square test was used to observe the association.

### Ethical Considerations

The study complied with the Declaration of Helsinki and was approved by the Ethical Review Committee, Department of Public Health, Northern University Bangladesh, Dhaka, Bangladesh (memo no. NUB/DPH/EC/2020/05). Participation of the respondents was anonymous and voluntary. Informed consent was sought from the respondents at the beginning point of the survey.

## Results

### Respondent characteristics

Among the total of 199 respondent mass media professionals, 155 (77.9%) were male with a male-female ratio of 3.5:1 and age range was 24 to 48 years with mean (±SD) age 33.59 (±4.67) years **(Table 1)**.

**Table 1:** Characteristics of the respondents (n=199)

|  |  |
| --- | --- |
| **Age (years)** | **Range: 24-48**<br>**Mean (±SD): 33.59 (±4.67)** |
| **Sex** | Male: 155 (77.9%)<br>Female: 44(22.1%)<br>Female Male ratio: 3.5:1 |
| **Existing comorbidities\*** | Absent: 122 (65.8%)<br>Present: 68 (34.2%)<br>- Asthma (27.05%)<br>- Hypertension (9.05%)<br>- Diabetes Mellitus (9.02%)<br>- Obesity (1.64%)<br>- Solid tumor malignancy (0.82%)<br>- Any rheumatic diseases (0.82%)<br>- History of bone marrow/stem cell transplant (0.82%). |
| **Symptoms\*** | Asymptomatic: 77 (38.7%)<br>Symptomatic: 122 (61.3%)<br>- Cough (77.87%)<br>- Fever (72.13%)<br>- Fatigue (53.28%)<br>- Headache (45.08%)<br>- Sore throat (55.74%)<br>- Myalgia (40.16%)<br>- Anosmia (30.33%) |

\*Percentage >100% due to Multiple Responses

Out of total respondents, 155 (57.8%) were found as never smoked any types of tobacco products, and 65.8% (n=122/199) didn’t have any kind of chronic medical conditions (comorbidities). Among the respondents, most were working in electronic media (69.3%, n = 138/199), followed by print media (16.6%, n=33/199) and online media (14.1%, n=28/199). Total 61 (30.7%) mass media professionals worked only in indoor office areas as editors, graphic designers, and make-up artists. But the majority (69.3%, n=138/199) were at multiple workplaces exposure (indoor office area, indoor studio, and outdoor video shooting area) as a reporter (n=65/138), news presenter (n=50/138), and video journalist (n=23/138). Mostly (89.9%, n = 179/199) were exposed in social gatherings of outside workplace settings like dynein restaurant, use public transport, and joined in gathering >10 people **(Table 2)**.

**Table 2:** Characteristics of the respondents according to their status of COVID-19 (n=199)

| Characteristics | COVID-19 status |  |  |  |
| --- | --- | --- | --- | --- |
|  | Number of participants, n (%) | Test Positive, n (%) | Suspected, Test negative and Healthy, n (%) | p-value (≤0.05) |
| **Age group** |  |  |  |  |
| ≤34 | 131 (65.8) | 41 (20.6) | 90 (45.2) | 0.01* |
| >34 | 68 (34.2) | 37 (18.6) | 31 (15.6) |  |
| **Sex** |  |  |  |  |
| Male | 155 (77.9) | 57 (28.6) | 98 (49.2) | 0.19 |
| Female | 44 (22.1) | 21 (10.6) | 23 (11.6) |  |
| **Tobacco smoking status** |  |  |  |  |
| Current | 49 (24.6) | 23 (11.6) | 26 (13.1) | 0.31 |
| Former | 35 (17.6) | 15 (7.5) | 20 (10.1) |  |
| Never | 115 (57.8) | 40 (20.1) | 75 (37.7) |  |
| **Existing Co morbidities** |  |  |  |  |
| Yes | 68 (34.2) | 39 (19.6) | 29 (14.6) | 0.01* |
| No | 131 (65.8) | 39 (19.6) | 92 (46.2) |  |
| **Types of media setting** |  |  |  |  |
| Print media | 33 (16.6) | 8 (4) | 25 (12.6) | 0.01* |
| Electronic media | 138 (69.3) | 66 (33.2) | 72 (36.2) |  |
| Online media | 28 (14.1) | 4 (2) | 24 (12.1) |  |
| **Types of job role in work setting** |  |  |  |  |
| Reporter | 65 (32.7) | 25 (12.6) | 40 (20.1) |  |
| Video Journalist | 23 (11.6) | 10 (5) | 13 (6.5) |  |
| News presenter | 50 (25.1) | 16 (8) | 34 (17.1) |  |
| Others (Editorial job, Graphic designer, Makeup artists and others) | 61 (30.7) | 27 (13.6) | 34 (17.1) |  |
| **Types of mask used at work place** |  |  |  |  |
| No respiratory protection | 9 (4.5) | 3 (1.5) | 6 (3) | 0.82 |
| Home-made/mask made by cloth | 14 (7.0) | 4 (2.0) | 10 (5.0) |  |
| New medical/surgical mask | 48 (24.1) | 20 (10.1) | 28 (14.1) |  |
| Re-used medical/surgical mask | 35 (17.6) | 12 (6.0) | 23 (11.6) |  |
| Respirator mask ( N95, FFP2, FFP3) | 93 (46.7) | 39 (19.6) | 54 (27.1) |  |
| **Symptoms** |  |  |  |  |
| Symptomatic | 122 (61.3) | 74 (37.2) | 48 (24.1) | 0.01* |
| Asymptomatic | 77 (38.7) | 4 (2) | 73 (36.7) |  |
| **Seeking maximum medical care support** |  |  |  |  |
| Over telephone/ via telemedicine with self-management at home | 108 (54.3) | 60 (30.2) | 48 (24.1) |  |
| Needed hospitalization | 15 (7.5) | 14 (7) | 1 (0.5) | 0.01* |
| No treatment needed/ Healthy respondent | 76 (38.2) | 4 (2) | 72 (36.2) |  |
| **Contact tracing of COVID-19** |  |  |  |  |
| Colleagues | 72 (36.2) | 33 (16.6) | 39 (19.6) | 0.09 |
| Guests/ Interviewees | 21 (10.6) | 11 (5.5) | 10 (5) |  |
| Friends & family | 8 (4) | 4 (2) | 4 (2) |  |
| Not Identified | 98 (49.2) | 30 (15.1) | 68 (34.2) |  |
| **Types of workplace exposure** |  |  |  |  |
| Only Indoor office area | 61 (30.7) | 22 (11.1) | 39 (19.6) |  |
| Multiple exposure (In studio/Indoor video shooting area, In outdoor video shooting area, interview/news presentation) | 138 (69.3) | 56 (28.1) | 82 (41.2) | 0.55 |
| **Exposure to outside workplace** |  |  |  |  |
| Not exposed to social gathering outside workplace | 20 (10.1) | 10 (5) | 10 (5) |  |
| Exposed to social gatherings (Used Public Transport, dyne in restaurant and joined in gathering >10 people) | 179 (89.9) | 68 (34.2) | 111 (55.8) | 0.29 |

Data are presented as frequency (n), percentage (%); \*Statistical significance at p value $\leq 0.05$ . Chi-square test was used to observe the association.

### Identified COVID-19 infection among media professionals

Out of total respondents, 39.2% (n=78/199) were revealed as test positive in contrast to 16.1% who had experienced an illness suspicious for COVID-19, but test (RT PCR) was negative, 6% were suspected/symptomatic but didn’t do the test, and 38.7% were reported healthy but worked in the same work setting during the pandemic **(Fig 1)**.

**Fig1:**
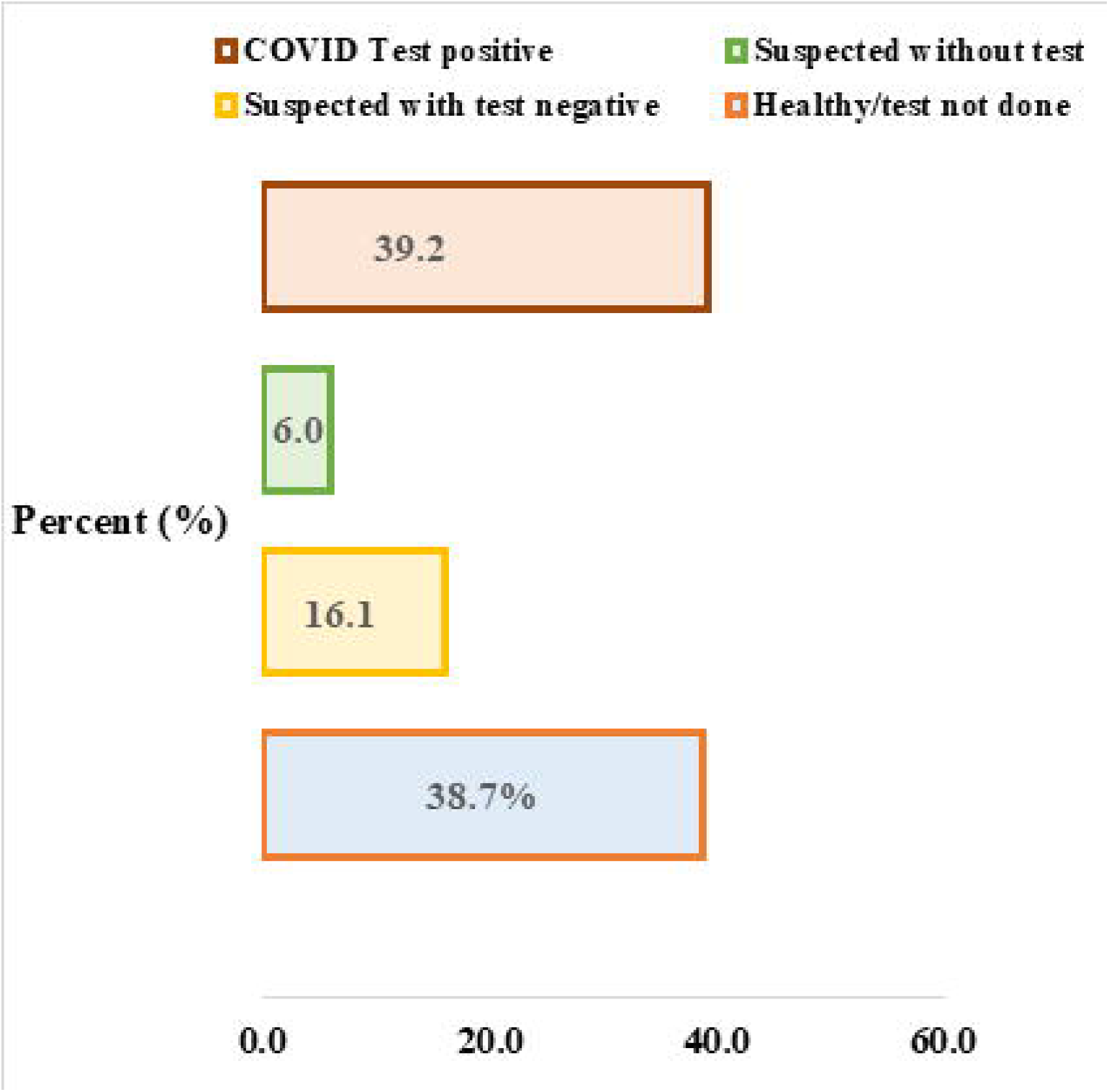
COVID-19 infection status among mass media professionals (n= 199)

Results of multivariate (cross table) and binary logistic regression analyses are shown in **Tables 2 and 3**. The study showed that the COVID-19 test was significantly more positive (OR/P= 0.38/0.01, AOR/P= 0.43/0.04) among the older age group (>34 years), and female respondents were identified more infected than male (OR= 0.64). The study didn’t find any association between tobacco smoking and COVID-19 infection, but respondents who had existing co-morbidities (OR/P= 3.17/0.01) were found more infected **(Table 3)**.

**Table 3.**
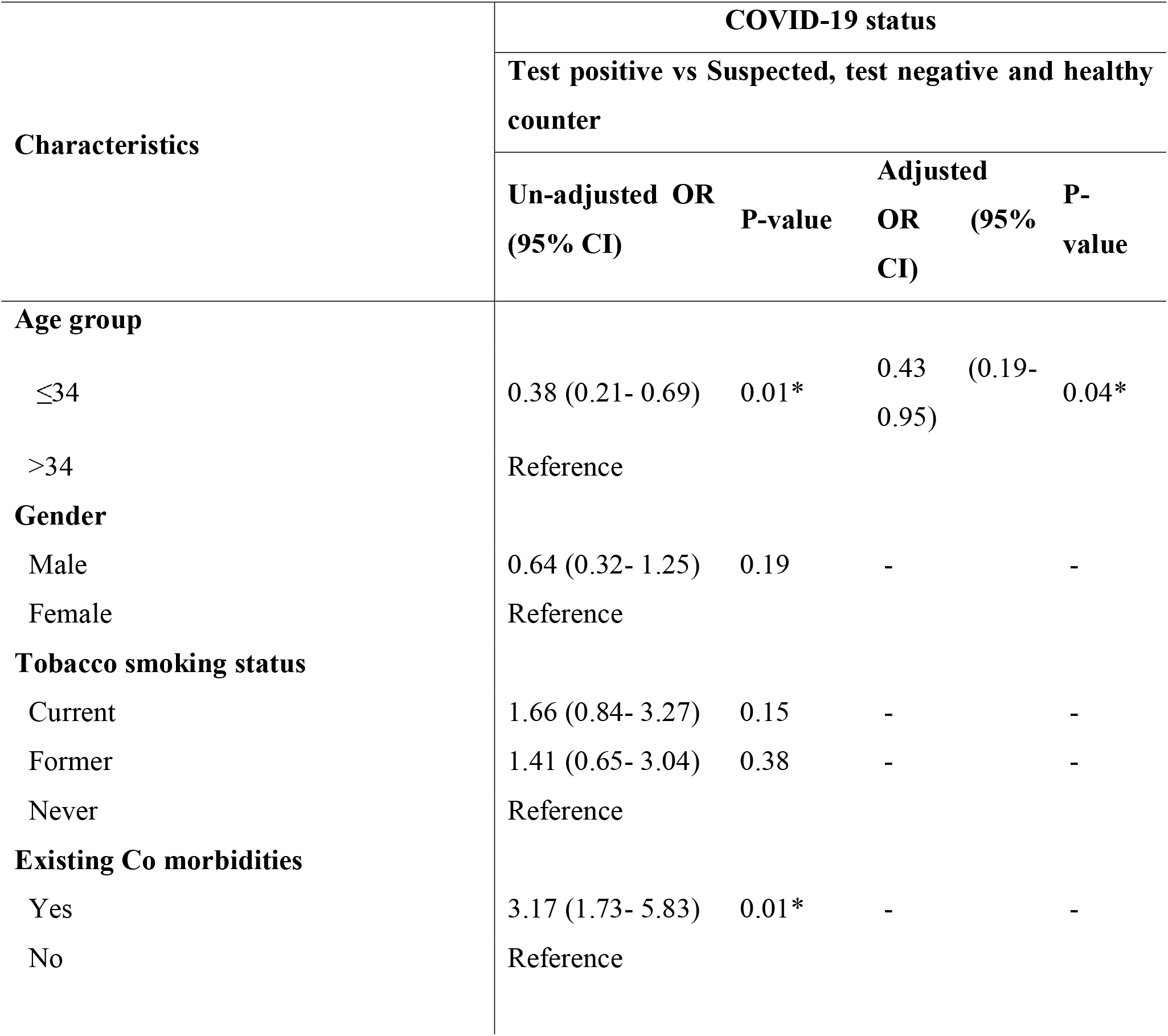
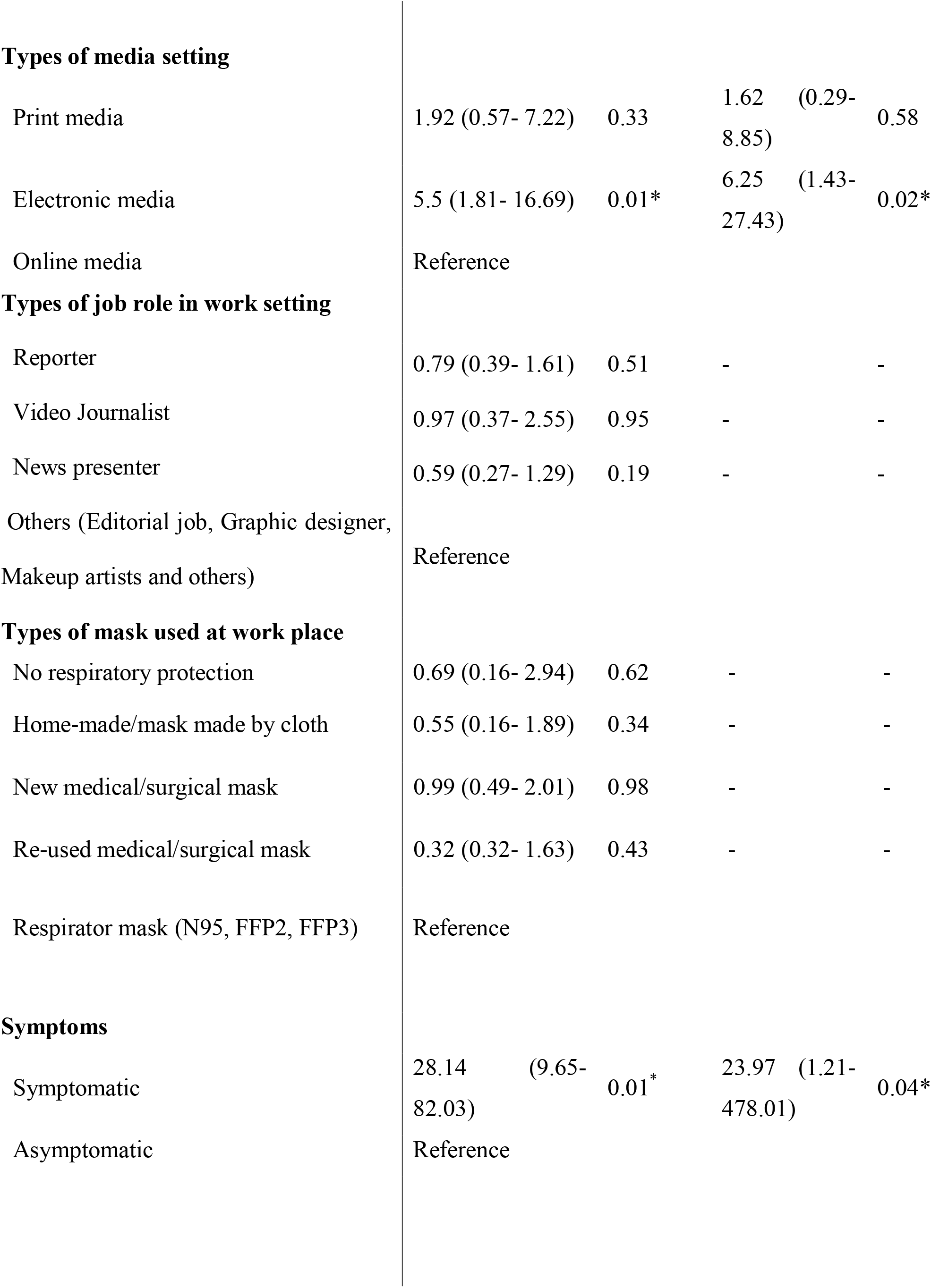
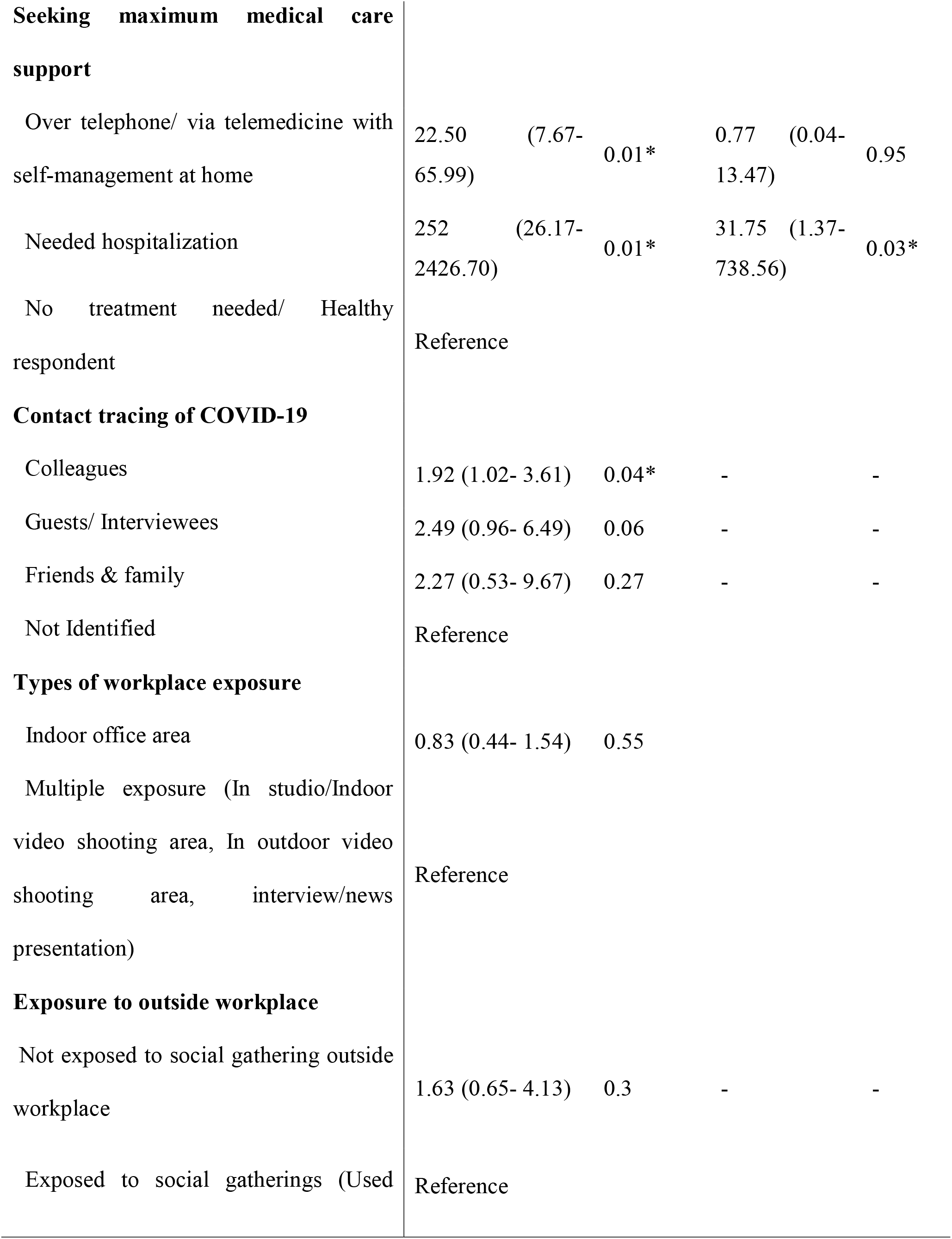
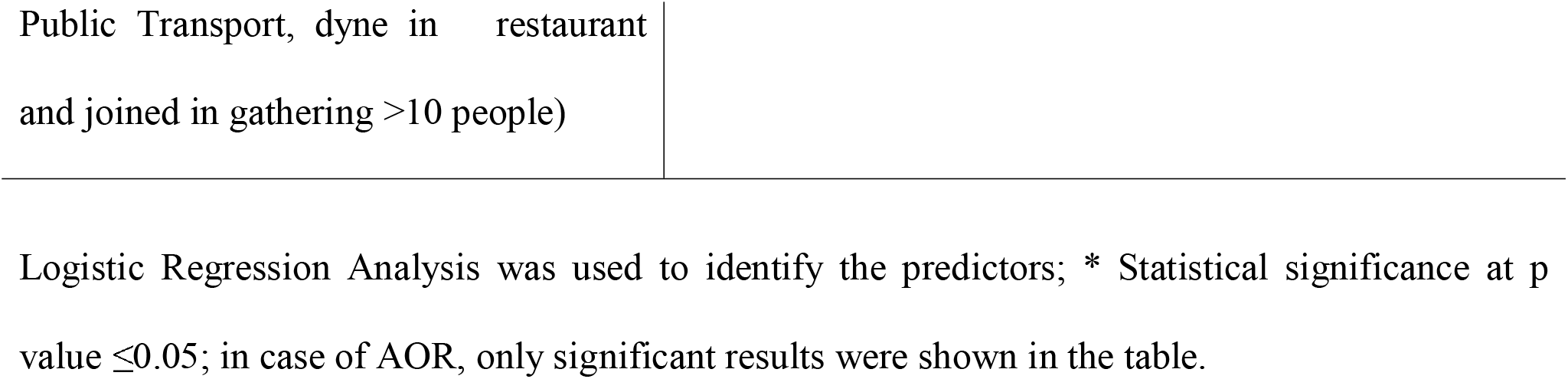
Association of COVID-19 infection and participants characteristics (n= 199)

| Characteristics | COVID-19 status |  |  |  |
| --- | --- | --- | --- | --- |
|  | Test positive vs Suspected, test negative and healthy counter |  |  |  |
|  | Un-adjusted OR<br>(95% CI) | P-value | Adjusted OR<br>(95% CI) | P-value |
| **Age group** |  |  |  |  |
| ≤34 | 0.38 (0.21- 0.69) | 0.01* | 0.43 (0.19- 0.95) | 0.04* |
| >34 | Reference |  |  |  |
| **Gender** |  |  |  |  |
| Male | 0.64 (0.32- 1.25) | 0.19 | - | - |
| Female | Reference |  |  |  |
| **Tobacco smoking status** |  |  |  |  |
| Current | 1.66 (0.84- 3.27) | 0.15 | - | - |
| Former | 1.41 (0.65- 3.04) | 0.38 | - | - |
| Never | Reference |  |  |  |
| **Existing Co morbidities** |  |  |  |  |
| Yes | 3.17 (1.73- 5.83) | 0.01* | - | - |
| No | Reference |  |  |  |
| **Types of media setting** |  |  |  |  |
| Print media | 1.92 (0.57- 7.22) | 0.33 | 1.62 (0.29- 8.85) | 0.58 |
| Electronic media | 5.5 (1.81- 16.69) | 0.01* | 6.25 (1.43- 27.43) | 0.02* |
| Online media | Reference |  |  |  |
| **Types of job role in work setting** |  |  |  |  |
| Reporter | 0.79 (0.39- 1.61) | 0.51 | - | - |
| Video Journalist | 0.97 (0.37- 2.55) | 0.95 | - | - |
| News presenter | 0.59 (0.27- 1.29) | 0.19 | - | - |
| Others (Editorial job, Graphic designer, Makeup artists and others) | Reference |  |  |  |
| **Types of mask used at work place** |  |  |  |  |
| No respiratory protection | 0.69 (0.16- 2.94) | 0.62 | - | - |
| Home-made/mask made by cloth | 0.55 (0.16- 1.89) | 0.34 | - | - |
| New medical/surgical mask | 0.99 (0.49- 2.01) | 0.98 | - | - |
| Re-used medical/surgical mask | 0.32 (0.32- 1.63) | 0.43 | - | - |
| Respirator mask (N95, FFP2, FFP3) | Reference |  |  |  |
| **Symptoms** |  |  |  |  |
| Symptomatic | 28.14 (9.65- 82.03) | 0.01* | 23.97 (1.21- 478.01) | 0.04* |
| Asymptomatic | Reference |  |  |  |
| **Seeking maximum medical care support** |  |  |  |  |
| Over telephone/ via telemedicine with self-management at home | 22.50 (7.67- 65.99) | 0.01* | 0.77 (0.04- 13.47) | 0.95 |
| Needed hospitalization | 252 (26.17- 2426.70) | 0.01* | 31.75 (1.37- 738.56) | 0.03* |
| No treatment needed/ Healthy respondent | Reference |  |  |  |
| **Contact tracing of COVID-19** |  |  |  |  |
| Colleagues | 1.92 (1.02- 3.61) | 0.04* | - | - |
| Guests/ Interviewees | 2.49 (0.96- 6.49) | 0.06 | - | - |
| Friends & family | 2.27 (0.53- 9.67) | 0.27 | - | - |
| Not Identified | Reference |  |  |  |
| **Types of workplace exposure** |  |  |  |  |
| Indoor office area | 0.83 (0.44- 1.54) | 0.55 |  |  |
| Multiple exposure (In studio/Indoor video shooting area, In outdoor video shooting area, interview/news presentation) | Reference |  |  |  |
| **Exposure to outside workplace** |  |  |  |  |
| Not exposed to social gathering outside workplace | 1.63 (0.65- 4.13) | 0.3 | - | - |
| Exposed to social gatherings (Used | Reference |  |  |  |

|  |
| --- |
| Public Transport, dyne in restaurant<br>and joined in gathering >10 people) |

Logistic Regression Analysis was used to identify the predictors; \* Statistical significance at p value $\leq 0.05$ ; in case of AOR, only significant results were shown in the table.

The reported existing co-morbidities were Asthma (27.05%), Hypertension (9.05%), Diabetes Mellitus (9.02%), Obesity (1.64%), Solid tumor malignancy (0.82%), any rheumatic diseases (0.82%) and History of bone marrow/stem cell transplant (0.82%) (table 1). Most of the test positive mass media professionals were symptomatic (OR/P= 28.14/0.01, AOR/P= 23.97/0.04) (table 3). Cough (77.87%) was the most common symptoms reported by respondents followed by fever (72.13%), fatigue (53.28%), headache (45.08%), sore throat (55.74%), myalgia (40.16%) and anosmia (30.33%) **(Table1)**.

Approximately half (n=108/199, 54.3%) of the total respondents sought medical attention via telemedicine or consultation with a doctor over the telephone with self-management at home, and only 7.5% (n=15/199) required hospitalization **(Table 2)**. Most of the test positive mass media professionals also reported telemedicine or consultation with a doctor over the telephone (OR/P= 22.50/0.01) as their required highest medical care support **(Table 3)**.

### Types of job role and work place exposure of mass media professionals

Respondents who worked in electronic media reported significantly more COVID-19 infection (adjusted odds ratio, AOR= 6.25; 95% Confidence interval: Lower limit 1.43, upper limit 27.43; P =0.02) than other groups **(Table 3)**. Out of total respondents, 30.7% (n=61/199) was working only in indoor office area (editorial job, graphic designer, makeup artist), and among them, 13.6% (n=27/199) reported test positive. On the other hand, 69.3% (n=138/199) of participants had exposure to various work locations, including indoor office areas, indoor & outdoor video shooting areas, and 28.1% (n=56/199) reported test positive. Out of total mass media professionals who worked in multiple locations, reporters had the highest number of test-positive infection (n=25/199, 12.6%) followed by news presenter (n=16/199, 8%) and video journalist (n=10/199, 5%) (Table 2). However, the study didn’t find a significant relationship between the job role or workplace exposure and COVID-19 infection. Though almost half of the participants (n=98/199, 49.2%) didn’t identify the contact of acquiring infection **(Table 2)** furthermore, infected colleagues (OR/P= 1.92/0.04) were reported as significant contact by the test positive mass media professionals **(Table 3)**.

### Exposures outside the mass media professional’s workplace

Results showed that most mass media professionals (n=179, 89.9%) were exposed to social gatherings like dyne in restaurants, used public transport, and joined in gathering>10 people. However, among the 10.1% (n=20/199) of respondents who didn’t have exposure to social gatherings, 5% (n=10/199) reported test positive for COVID-19 (table 2). The study didn’t find a significant association between the outside workplace exposure and COVID-19 infection of mass media professionals (OR/P= 1.63/0.3) **(Table 3)**.

### Uses of different type of mask and respirator

Total 46.7% (n=93/199) mass media professionals reported that they used respirator mask (N95/FFP2/FFP3) while on duty and others used new medical mask (n=48/199, 24%), reused medical mask (n=35/199, 17.6%) and home-made/made by cloth mask (n=14/199, 7%). The study result showed that reused medical masks, homemade or masks made by cloth were not significantly (p=0.82) associated with mass media professionals’ infection **(Table 2)**.

## Discussion

Since the emergence of the COVID-19 pandemic, studies [15, 16] among front-line employees primarily included healthcare workers. But other groups of front-line employees are at risk of being infected during work [17, 18]. We intended to capture comprehensive data related to mass media professions infection and risk of COVID-19. The occupational exposure of mass media professionals is different from other professions. Due to their close interaction with the community, gathering more than 10 people and using public transport is not always extra-occupational exposures and some exposures are inevitable. Thus, media professionals are a high-risk group and potential to being and transmit infection. So, essential measures and attention to these frontlines therefore warranted.

This study found 39.2% of mass-media professionals in Bangladesh had test positive COVID-19 infection. The result suggested that respondents who worked in electronic media had more COVID-19 infection. Reporters had multiple workplace exposures, and the study revealed that they are the vulnerable group for being infected with COVID-19. Conversely, it was anticipated that media professionals who worked only in indoor office areas would be at very low risk for acquiring infection at the workplace, but it was therefore surprising that 13.6% of respondents who were working only in the indoor office area revealed test positive COVID-19 infection. Furthermore, infected colleagues were reported as significant contact by the test positive mass media professionals. On the other hand, 16.1% of mass media professionals were reported to test negative but symptomatic. According to Arevalo-Rodriguez I et al. [19], up to 54% of COVID-19 patients may have an initial false-negative RT-PCR. Protective behavior, including maintaining physical distancing while at work and not working while ill, remains pertinent to mass media professionals.

The study showed that wearing a respirator mask while at work didn’t significantly protect mass media professionals from COVID-19 infection. However, using a medical mask, homemade or cloth mask, was not significantly associated with mass media professionals’ infection. A meta-analysis of randomized trials by Bartoszko JJ et al [20] also revealed that medical masks and respirator masks offer similar protection from COVID-19 infection in healthcare workers.

This study has several strengths. First, it is the 1^st^ reported study revealing COVID-19 status among media professionals. Second, we used an online survey link to rapidly obtain prospective data from different country areas within an ongoing pandemic. Third, we obtained information from participants who did not do the test or have negative RT-PCR reports, which assessed risk factors with minimal recall bias. Fourth, we also recorded initial onset, minimal symptoms, or asymptomatic cases, minimizing biases related to capturing only severe hospitalization records.

## Conclusion

This study suggested that 39.2% of mass-media professionals in Bangladesh had tested positive COVID-19 infection, and risk was significantly high among those working in electronic media. The study underscores the need to wear respirator masks by mass media professionals while at work, as respirator mask, medical mask, and homemade/Cloth-made mask revealed offer almost similar protection from COVID-19 infection in mass media professionals. Infected colleagues were reported as significant contact for acquiring infection. Protective behavior, including maintain physical distancing while at work and not working while ill, remains pertinent to mass media professionals. Closer scrutiny of infection control measures surrounding them should be an essential concern and need for paying attention as they largely huddled in the community while discharging their duties. Findings of this study will aid the policy maker and public health authorities during the COVID-19 pandemic to make proper implementation measures.

## Limitations

We acknowledge our several limitations. First, our findings are based on the self-report of respondents. As self-selected into study participation, so selection or collider bias is possible. Second, as all enrolled healthy/asymptomatic media professionals did not do the test (RT-PCR), some crossover of asymptomatic infection is possible for the healthy media professionals. It may reduce the odds ratio. Third, additional detailed information on whether the participant’s family belonged to a high-risk group or exposure would have been pertinent to the study. Finally, the possibility of unmeasured confounders is always present, although our primary conclusions were robust to several sensitivity and secondary analyses. Further follow-up of these observational findings is needed.

## Data Availability

Data and materials supporting our findings in the manuscript will be shared on request.

## Acknowledgments

The author acknowledges the support of Md. Farhad Mia in process of the web link formation and data entry. Additionally, thanks to all participants who gave their valuable time for complete the self-administered online survey.

## Declarations

### Funding

This was a self-funded study with no external funding support for any aspect of this study

### Use of the manuscript or data

this manuscript/data was not used in any form of presentation or used for any form of publication.

### Conflict of interest

The authors declared none competing interest.

### Consent of Publication

All authors had full access to the full data in the study and accept responsibility to submit for publication.

### Author contributions

- **ST:** Study concept & design, literature search, data collection, data interpretation, writing, study supervision and critical revision
- **BB:** Study design, data analysis, data interpretation, writing
- **NA:** Data analysis, data interpretation, writing
- **SMH:** Writing review and editing
- **RAM:** Writing review and editing
- **MRA:** Writing review and editing

### Availability of data and material

Data and materials supporting our findings in the manuscript will be shared on request.

## Notes

### Competing Interest Statement

The authors have declared no competing interest.

